# Evaluation of the protective efficacy of Olyset®Plus ceiling nets for reduction of malaria incidence in children in Homa Bay County, Kenya: a cluster-randomized controlled study protocol

**DOI:** 10.1101/2024.04.20.24306116

**Authors:** Yura K Ko, Wataru Kagaya, Protus Omondi, Kelvin B. Musyoka, Takatsugu Okai, Chim W. Chan, James Kongere, Victor Opiyo, Jared Oginga, Samuel M. Mbugua, Bernard N. Kanoi, Mariko Kanamori, Daisuke Yoneoka, Kenya National Bureau of Statistics (KNBS), Kibor Keitany, Elijah Songok, Gordon Okomo, Noboru Minakawa, Jesse Gitaka, Akira Kaneko

## Abstract

**Introduction:** Malaria is still a major health problem in sub-Saharan Africa, where 98% of global malaria mortality occurs. In addition, the spread of *Plasmodium falciparum* with partial artemisinin resistance in East Africa and beyond is a great concern. The establishment of more effective vector control, in addition to the current long-lasting insecticide-treated net (LLIN) distribution program, is an urgent task in these areas. One novel vector control candidate is the Olyset®Plus ceiling nets which can overcome the problems of variations in net use behaviors and metabolic resistance to insecticide in vectors. Our preliminary study suggests the protective efficacy and high acceptability of this tool. With this proposed second trial, we aim to evaluate the impact of this tool in a different eco-epidemiological setting in the lake endemic region of Kenya.

**Methods:** A cluster randomized controlled trial is designed to evaluate the impact of Olyset®Plus ceiling nets in Ndhiwa Sub-County, Homa Bay County, Kenya. A total of 44 clusters will be randomly assigned in a 1:1 ratio to the intervention group (Olyset®Plus ceiling nets) and the control group. The assignment will be accomplished through covariate-constrained randomization of clusters. For the primary outcome of clinical malaria incidence, 38 children from each cluster will be enrolled in a cohort and followed for 18 months. We will also evaluate the effects of the intervention on entomological indicators as well as its acceptance by communities and cost-effectiveness.

**Ethics and dissemination:** Ethics approval was provided by the Mount Kenya University Institutional Scientific Ethics Review Committee. Study results will be shared with study participants and communities, the Homa Bay County Government and the Kenya National Malaria Control Programme. Results will also be disseminated through publications, conferences and workshops to help the development of novel malaria control strategies in other malaria-endemic countries.

**Trial registration:** UMIN000053873

**Administrative information:** 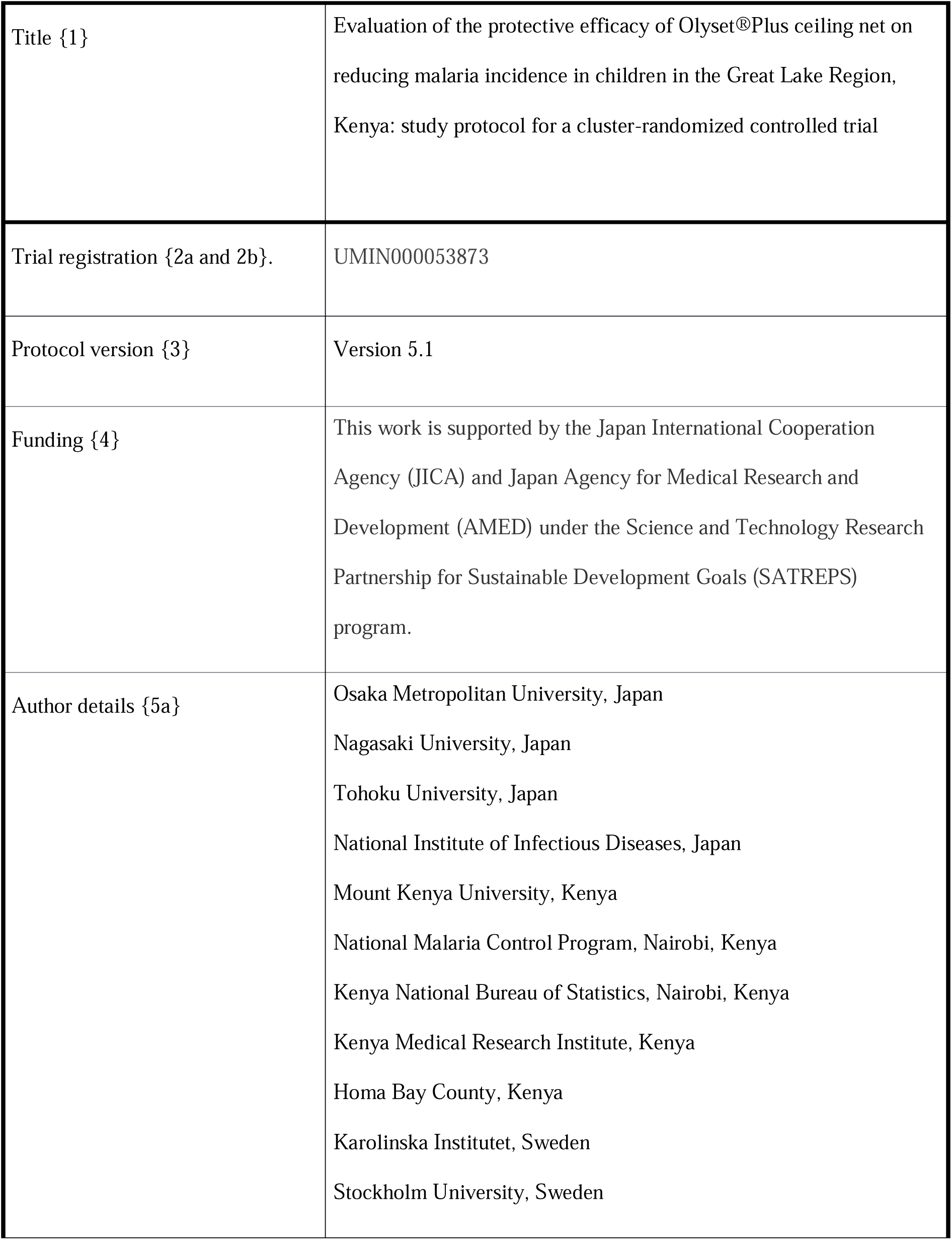

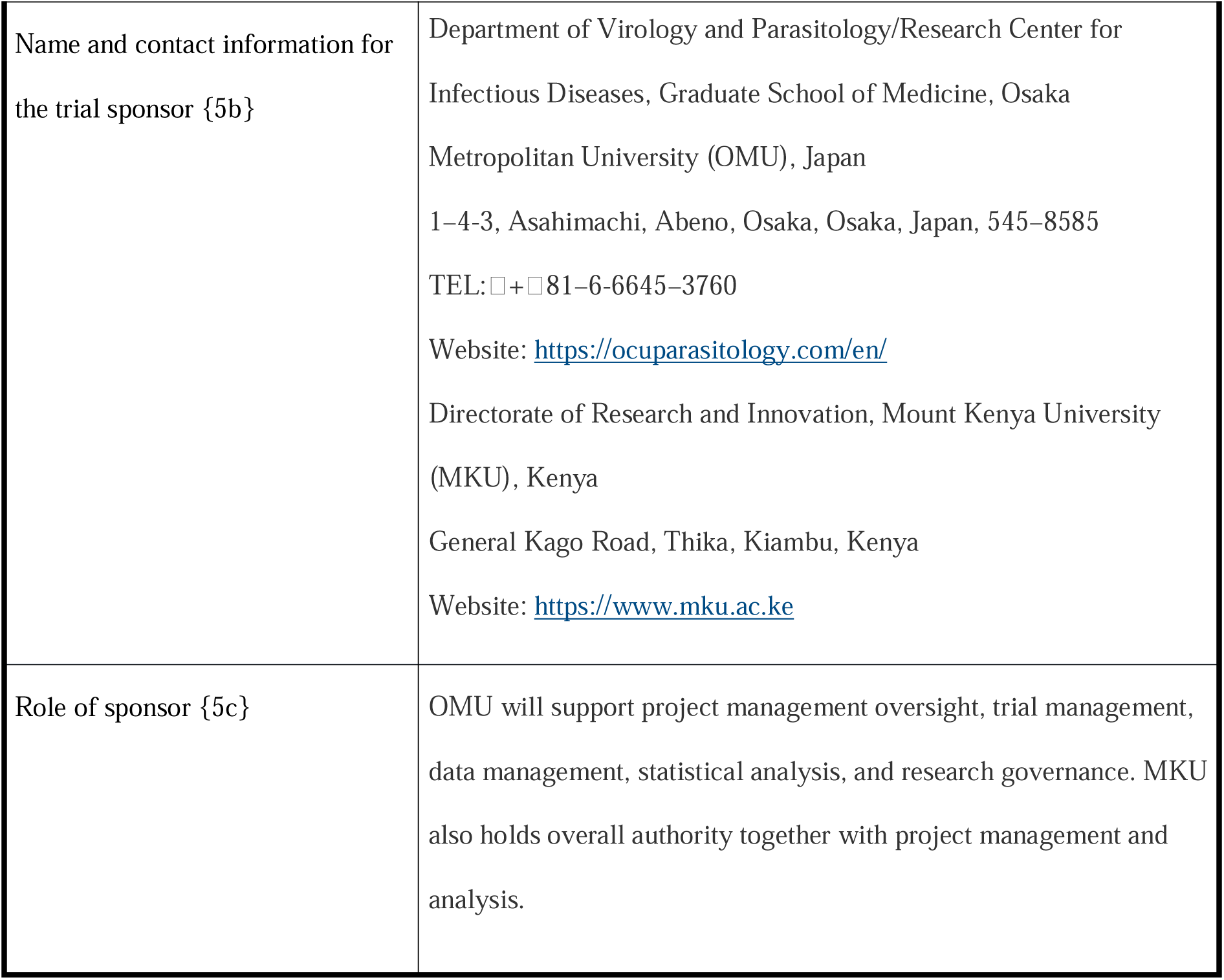

**Strength and limitations of this study:**

- This study is a cluster-randomized controlled trial (CRCT) to evaluate the efficacy of the Olyset®Plus ceiling net as a novel vector control tool and a complement to current malaria control tools in sub-Saharan Africa.
- This marks the second CRCT of the Olyset®Plus ceiling net intervention in the lake endemic region of Kenya, expanding the evidence base to a different eco-epidemiological setting from the previous CRCT, where promising results were observed on Mfangano Island.
- Collaboration with local Kenyan institutions such as the Kenya National Bureau of Statistics (KNBS), the National Malaria Control Programme (NMCP), the Kenya Medical Research Institute (KEMRI), and Homa Bay County from the research planning stage is one of the strengths of this trial, allowing for a seamless transition from research implementation in the field to policy development.
- One of the anticipated limitations is the possible contamination between intervention and control clusters because we will not set a buffer zone due to the geographical proximity of each cluster. We will try to account for such contamination effects by integrating spatial data into our statistical model.

## Introduction

### Background and rationale {6a}

Malaria is still a major health problem, particularly in sub-Saharan Africa, where 98% of global malaria mortality occurs [1]. Although the morbidity and mortality of malaria declined from the 2000s to 2015 owing to many investments and interventions, such as long-lasting insecticide-treated nets (LLINs), malaria rapid diagnostic tests (RDTs), and artemisinin-based combination therapies (ACTs), progress has stalled since 2015. Moreover, the spread of *Plasmodium falciparum* partially resistant to ACT in Africa is an enormous concern. Currently five African countries, Rwanda [3], Uganda [4], Eritrea [5], Ethiopia [6], and the United Republic of Tanzania [2] have reported delayed clearance of *P. falciparum* after treatment with ACTs. Kenya’s proximity to these countries highlights the urgent need to establish effective vector control, in addition to maintaining antimalarial drug efficacy and strengthening resistance surveillance.

Among several vector control measures, LLIN is the most widely adopted tool to prevent mosquito bites and interrupt malaria transmission. However, suboptimal uses of LLIN are key factors in reducing the impact of LLIN on the malaria burden. In the Lake Victoria basin, alternative uses of LLIN for fishing and protecting crops and chicks are well-known local behaviors [7,8]. In fact, in many areas including our study sites in Homa Bay County, Kenya, malaria prevalence remains high despite widespread distribution of LLINs and their periodic replacements for more than a decade. This suggests that LLIN alone is insufficient to interrupt malaria transmission in this region.

Recently, we have proposed a novel vector control tool that covers the ceiling and the gap between the ceiling and the walls of residential structures with co-formulated pyrethroid and piperonyl butoxide (PBO) bed net material, called the Olyset®Plus ceiling net. The benefit of installing the Olyset®Plus ceiling net in addition to conventional LLINs is detailed elsewhere [9]. Briefly, the Olyset®Plus ceiling net provides a combination of physical and chemical protection against mosquitoes which seek human bloodmeal in the house. Furthermore, the ceiling net is semi-permanently installed and requires no further action from end users, thus its protective efficacy is consistently extended to all who sleep in the house and less affected by the variation in conventional LLIN use.

The aim of this study is to evaluate the efficacy, acceptability, and cost-effectiveness of Olyset®Plus ceiling nets on malaria morbidity and transmission in the Lake Victoria basin of Kenya. Preliminary data from our previous study on Mfangano Island in Lake Victoria [9] suggest a substantial reduction in malaria prevalence among school children and high community acceptance of this tool (unpublished data). With this proposed second trial, we aim to evaluate the impact of this tool in a different eco-epidemiological setting with relatively higher malaria transmission, more frequent human and vector movement, and synergistic impact from other interventions such as indoor residual spraying (IRS) and the RTS,S malaria vaccine. Since effective malaria controls need to be tailored to the local context, evidence of the effectiveness of Olyset®Plus ceiling nets from various transmission settings will increase the appeal of this intervention.

Furthermore, considering the recent increase in choices of malaria control tools and the necessity of combining various tools to maximize the impact of the malaria control program, it is important to understand the acceptability and cost-effectiveness of each intervention to guide its future deployment.

To achieve these objectives, our collaboration with local institutions including the Kenya National Bureau of Statistics (KNBS), the National Malaria Control Programme (NMCP), the Kenya Medical Research Institute (KEMRI), and Homa Bay County, started from the research planning stage. This collaboration is crucial to the seamless transition from field trial to expanded implementation and policy development.

### Objectives {7}

The study has four research domains: epidemiology, entomology, social aspects, and cost-effectiveness.

For the epidemiology domain, the primary objective is to determine the protective efficacy of Olyset®Plus ceiling net in reducing malaria clinical incidence in children 6 months to 14 years old over 18 months post-intervention. The secondary objectives are (1) to determine the protective efficacy of Olyset®Plus ceiling nets in reducing *Plasmodium* infection prevalence by PCR in all age groups at 6-, 12-, and 18-months post-intervention; (2) to determine the protective efficacy of Olyset®Plus ceiling nets against the time-to-first *Plasmodium* infection; (3) to determine the spillover effects of Olyset®Plus ceiling nets in reducing *Plasmodium* infection prevalence in all age groups at 6-, 12-, and 18-months post-intervention; and (4) to determine the protective efficacy of Olyset®Plus ceiling net in reducing *Plasmodium* infection incidence in children 6 months to 14 years old over 18 months post-intervention.

For the entomology domain, the primary objective is to evaluate the impact of Olyset®Plus ceiling nets on the mosquito density of the primary malaria vector species. The secondary objectives are (1) to determine the impact of Olyset®Plus ceiling nets on entomological inoculation rate (EIR) and (2) to determine the prevalence of knockdown resistance (*kdr*) mutations in vectors.

For the social aspects domain, the primary objective is to assess the determinants of social acceptability of the Olyset®Plus ceiling net in both the intervention and control arms. The secondary objectives are (1) to determine the feasibility of installing the Olyset®Plus ceiling and (2) to determine the appropriateness of fit of the Olyset®Plus ceiling net in the context of households in Ndhiwa Sub-County.

For the cost-effectiveness domain, the primary objective is to determine the incremental cost effectiveness ratios (ICERs) of adding the Olyset®Plus ceiling net to existing malaria control interventions under field trial conditions. The secondary objectives are (1) to establish the relative contribution to costs of the distinct programmatic elements and identify the inputs that contribute the most to overall costs, and (2) to estimate the potential cost of providing Olyset®Plus ceiling net at a larger scale over 3 and 5 years under operational scenarios.

### Trial design {8}

The study is an open-label, cluster-randomized controlled trial (CRCT) with 44 clusters evenly divided between the intervention and the control arms. Each cluster will include one or two villages and consist of at least 50 households. A baseline survey will be conducted to determine the pre-intervention *Plasmodium* prevalence and *Anopheles* density, and to collect demographic and socioeconomic data for covariate-constrained randomization of clusters. The baseline survey will be conducted one month before cluster randomization. The post-intervention follow-up period will be 18 months. For the evaluation of the primary objective, 38 children aged 6 months to 14 years from each cluster will be recruited and followed for 18 months as a cohort. Cross-sectional surveys will be conducted after 6, 12, and 18 months of the intervention targeting 50 individuals of all ages from each cluster to estimate the overall *Plasmodium* prevalence.

## Methods: Participants, interventions and outcomes

### Study setting {9}

#### Location and administrative structure

Ndhiwa (713.5 km²) is one of nine sub-counties in Homa Bay County in Kenya. The sub-county has seven administrative wards: Kanyamwa Kologi, Kanyamwa Kosewa, Kabuoch North, Kwabwai, Kanyadoto, Kanikela, and Kabuoch South/Pala. Based on the number of malaria cases reported in the Kenya Health Information System (KHIS), the accessibility of the site, and population size, we selected Kanyamwa Kologi Ward as the target area (Figure 1). Agriculture is the primary economic activity, with sugarcane as a main commercial crop. County residents also keep animals such as dairy cattle, beef cattle, sheep, goats, and poultry [10]. The ward experiences a long rainy season from March to June and a short rainy season from October to December. As of 2019, the mean annual precipitation is 228.64 mm and the mean annual temperature is 26.7□. The relative humidity remains elevated year-round, fluctuating between 75% to 85% [11].

**Figure 1:**
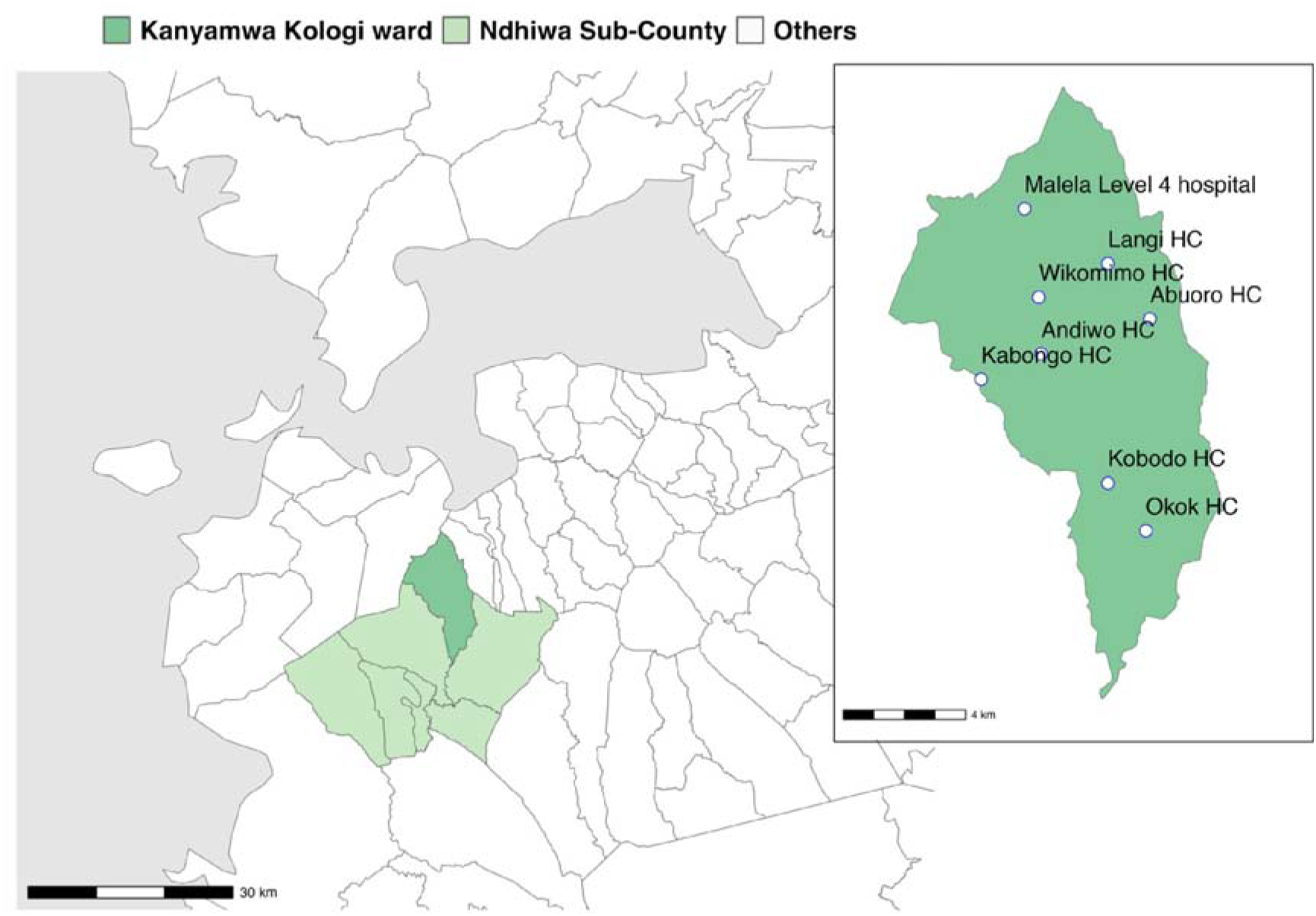
Map of the study area. Inset shows the locations of the level four hospital and seven health centers (HC) in Kanyamwa Kologi Ward.

#### Demographics

The population of Kanyamwa Kologi Ward is approximately 33,000 according to the 2019 national census [12]. The dominant ethnic group in the region is Luo and the primary languages are DhoLuo, Kiswahili, and English. There are 172 primary and 36 secondary schools in Ndhiwa Sub-County [13]. Within Kanyamwa Kologi Ward, there are 28 primary and 7 secondary schools.

#### Malaria epidemiology and control measures

Based on the KHIS, there were 429.1 and 457.2 confirmed malaria cases per 1,000 population in Ndhiwa Sub-County and Kanyamwa Kologi Ward, respectively, in 2023. The primary malaria vector in the sub-county is *Anopheles funestus*, which prefers feeding on humans. *An. arabiensis* is also an important malaria vector [14]. In Homa Bay County, LLINs have been distributed every three years since the early 2000s, and IRS and the RTS,S malaria vaccine have been piloted in several areas since 2018 and 2019, respectively [15]. Notably PBO-incorporated LLINs (Veeralin®LN, manufactured by VKA Polymers, Tamil Nadu, India) were distributed in late 2023. In Kanyamwa Kologi Ward, there is one level four hospital and seven health centers.

### Eligibility criteria {10}

As the ceiling nets are installed per structure, we set the inclusion criteria on a structural basis. The inclusion criteria for the installation of the ceiling nets are (1) residential structures with at least one permanent resident aged 18 years or older in the household, (2) informed consent provided by a resident in the household, and (3) house structure amenable to ceiling net installation in terms of size of the structure, presence of eave, ceiling board, and vertical beams, and material of the top part of the wall. The applicability of the ceiling net installation will be assessed by experienced field staff. The exclusion criteria are (1) vacant structure, confirmed by at least two visits by community health promoters (CHPs), (2) dwelling structure to be vacated or destroyed within the study period, (3) not applicable house structure for the ceiling net installation, and (4) non-residential structures (school, shop, kitchen, storage, and toilet). The inclusion criteria for the prospective cohort are (1) children aged 6 months to 12 years old at the time of enrolment, (2) living in the study area at the time of Olyset®Plus ceiling net installation, (3) having no plan to leave or stay outside the study area for an extended period (longer than one month) over the 18-month follow-up period, and (4) informed consent provided by the participants or the parent or legal guardian. The exclusion criterion is having severe chronic illnesses. The inclusion criteria for cross-sectional malaria surveys for all age groups are (1) living in the study area during the study period, and (2) informed consent being provided by the participants or the parent or legal guardian before each survey. The exclusion criteria are (1) having severe chronic illnesses and (2) pregnancy known at the time of the surveys.

### Who will take informed consent? {26a}

Written informed consent will be obtained by the study team members who fully understand the study protocol. After eligibility is confirmed, the study team members will present to the potential participant a document containing all relevant information about the study in Luo and English. If the participant cannot read, study information will be conveyed verbally in Luo, Kiswahili, and/or English by the study team members. The potential participant will have opportunities to ask any questions. Agreement to participate will be sought only after the participant indicates complete understanding of the study.

### Additional consent provisions for collection and use of participant data and biological specimens {26b}

The study information document for ceiling net installation contains the study overview. In addition, the documents for cross-sectional and cohort surveys contain details on collecting, storing, and using personal data and biological specimens during the study.

## Interventions

### Explanation for the choice of comparators {6b}

In Kenya, LLIN is the most widely used malaria preventive measure. The Division of National Malaria Programme coordinates free LLIN distribution, and the county governments deliver LLINs to residents in all endemic counties every three years. In Homa Bay County, the RTS,S malaria vaccine has been implemented since 2019. The primary purpose of this trial is to demonstrate the superiority in malaria prevention of adding Olyset®Plus ceiling nets to the standard malaria control program. Thus, in the control arm, no Olyset®Plus ceiling nets will be installed, but LLIN use and RTS,S immunization will be allowed in the control and intervention arms as the current best practice. There is no plan for new LLIN distribution during the study period.

### Intervention description {11a}

In the intervention arm, Olyset®Plus ceiling nets will be installed in all dwelling units where residents sleep, free of charge to the households. All participants will be encouraged to continue to use LLINs, distributed by the Homa Bay County government. In each intervention cluster, 1 CHP and 2 community volunteers will be recruited from the intervention cluster and another 1 CHP from the control cluster will join the team to enable future knowledge dissemination. The net installation team will be trained to install ceiling nets by skilled local research assistants who participated in previous trials. The head (or another adult) of the household eligible to a ceiling net will be notified at least 24 hours before the scheduled installation time. The cost of the ceiling nets and their installation will be covered by the research team. Details of the installation procedure are described in the previous study protocol [9]. Briefly, the ceiling net is a rectangular sheet of Olyset®Plus net with loops sewn along the diagonal seams. The loops are roped to the support beams under the roof and the edges of the net are stapled to the wall.

### Criteria for discontinuing or modifying allocated interventions {11b}

As the ceiling net is semi-permanently installed, the intervention will only be discontinued if the participant specifically requests the removal of the ceiling net by the study team. There will be no crossover from the control arm to the intervention arm during the follow-up period. Those who migrate between the arms or emigrate from the study areas will be dropped from the study follow-up.

### Strategies to improve adherence to interventions {11c}

Adherence to the intervention cohort in this study is defined as sleeping in houses with Olyset®Plus ceiling nets. Adherence is monitored indirectly by assessing the number of nights each participant spends outside their house during the bi-weekly interview. During each house visit, CHPs will visually inspect the condition of the ceiling nets. Any visible tear and damage to the ceiling net will be reported to the research team, who will assess the size and location of the damage and perform repair or replace the ceiling net if necessary.

### Relevant concomitant care permitted or prohibited during the trial {11d}

There is no specific concomitant care prohibited during the trial. All participants in both arms will continue to receive and use free LLIN and have access to standard medical care, including malaria testing by RDT, treatment with ACT and RTS,S malaria vaccination.

### Provisions for post-trial care {30}

All participants will be under the normal healthcare system in the study setting. No perceived health risks for the intended population are expected with the intervention. Our plan of continuous cross-sectional malaria surveillance after the study period allows us to monitor further parasite transmission in the population.

### Outcomes {12}

#### Epidemiological domain

The primary outcome will be symptomatic malaria case incidence, defined as axillary temperature of ≥37·5°C or a history of fever in the preceding 48 hours, and positive mRDT, in children aged 6 months to 14 years enrolled in the cohort, monitored with biweekly visit and passive case detection in the health facilities during an 18-month follow-up. The secondary outcomes will be (1) the prevalence of *Plasmodium* infections by PCR in all age groups at 6, 12, and 18 months post-intervention, (2) time to first infection defined as the number of days between the start of the intervention and the first PCR positive diagnosis in the cohort of children during the 18-month follow-up period, (3) spillover effect measured with the above-mentioned prevalence parameter, and (4) infection incidence by PCR in the prospective cohort of children aged 6 months to 14 years over 18 months.

#### Entomological domain

The primary outcome will be the density of the primary malaria vectors, species composition and sporozoite infection rates. Malaria vector density will be determined using CDC light trap, and species composition and sporozoite infection rates will be determined by microscopy and PCR. The secondary outcomes of the entomology domains will be (1) changes in EIR as a measure of malaria transmission and (2) prevalence of *kdr* mutations associated with insecticide resistance in *Anopheles* mosquitoes captured by light trap.

#### Social aspect domain

The primary outcome will be the percentage of households consenting to Olyset®Plus ceiling net installation when offered. In addition, we will include observations and discussions about individual attitudes toward the ceiling net. The secondary outcomes of the social aspect domain will be the percentage of the intact ceiling net, description of damaged net, and the impact on the living environment at 6, 12, and 18 months post-intervention.

#### Cost-effectiveness domain

The primary outcome will be the incremental cost-effectiveness of adding Olyset®Plus ceiling net to existing malaria control interventions under field trial conditions from the societal and provider perspectives. The secondary outcomes of the cost-effectiveness domains will be (1) the costs of the distinct programmatic elements and the inputs that contribute the most to overall costs, and (2) the cost of providing Olyset®Plus ceiling net at a larger scale over three and five years under operational scenarios.

### Participant timeline {13}

The schedule of trial activities is presented in Figure 2. The detail of each survey is described in Figure 3.

**Figure 2:**
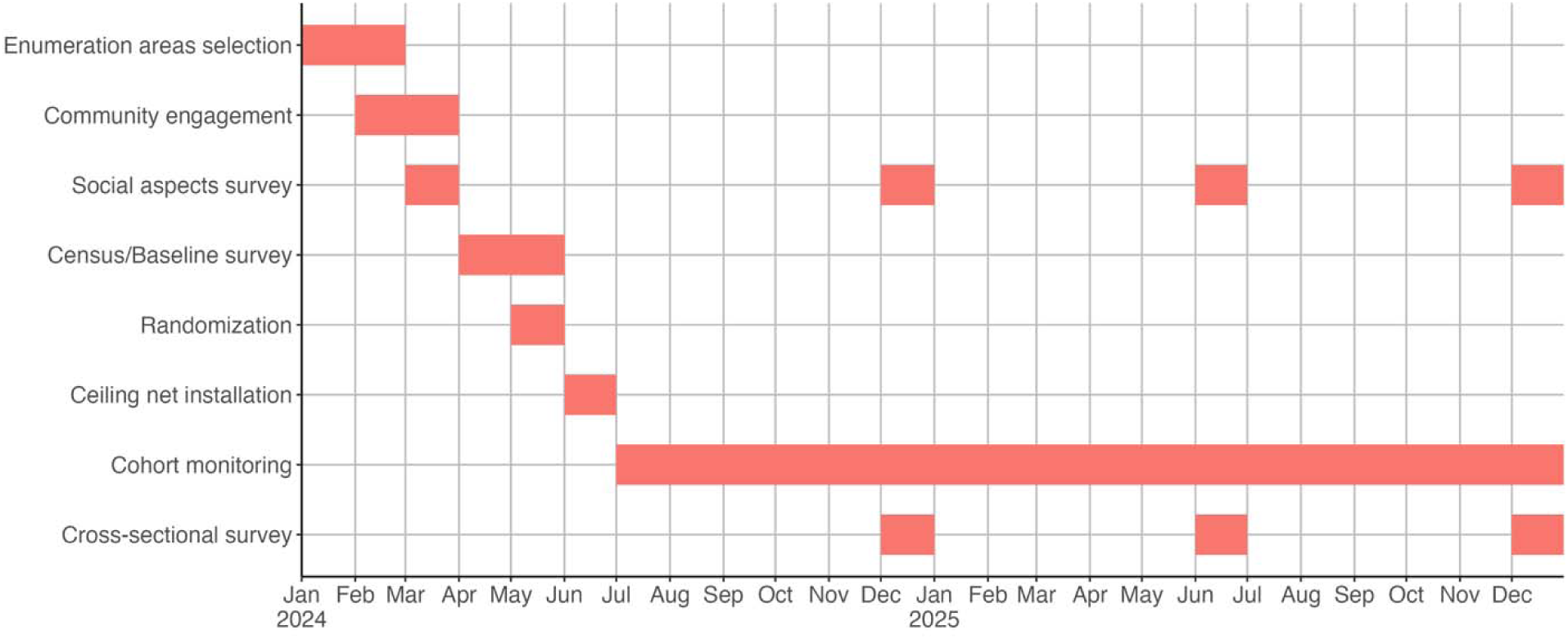
The schedule of trial activities.

**Figure 3:**
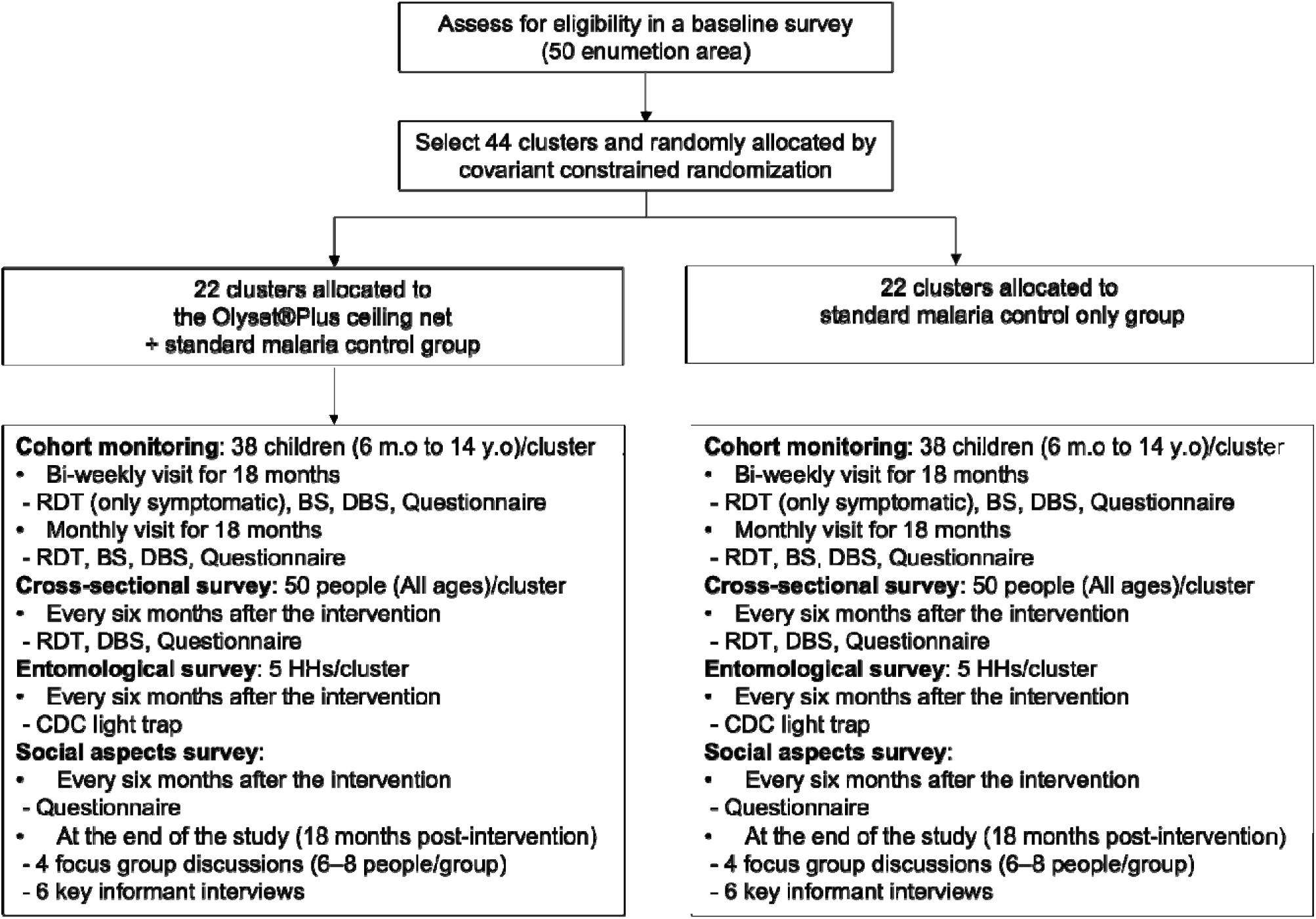
CONSORT flow diagram and the detail of each survey

### Sample size {14}

The sample size was calculated using the method of Hayes and Moulton [16]. All the sample sizes will be recalculated based on the baseline data, which will be collected about one month before the ceiling net installation.

#### Epidemiological survey

The following calculations were based on the historical data collected from the Lake Victoria region in Kenya with a clinical malaria incidence rate of 0.5 per person-year in children under 14 years old by RDT (unpublished data on Mfangano island), 40% parasite prevalence for all age groups by PCR and a between-cluster coefficient of variation (CV) in incidence rate of 0.24 in both groups. In the study site, RTS,S malaria vaccination began in 2019, with a mass distribution of PBO-incorporated LLINs at the end of 2023.

Therefore, the intervention effect is expected to be smaller than those in previous studies and is conservatively assumed to be 25%. Assuming 38 individuals per cluster to be followed for up to 18 months with 20 % loss-to-follow up rate, we require 22 clusters per arm to achieve 80% power to detect a significant incidence rate ratio of 0.75 (25% protective efficacy) at a two-sided type 1 error of 5%. With the number of clusters with 50 individuals per cluster, for the secondary outcome of malaria prevalence by PCR for all age groups, we would achieve 5% type 1 error and 80% power to detect 23.5 % relative reduction. We do not specify the sample size for identifying spillover effects because spillovers tend to have smaller effect sizes relative to total or overall effects, so typically larger sample sizes are required to detect them. Although our study may be underpowered to detect spillovers, we will report the results as an exploratory analysis.

#### Entomological survey

Based on the previous study, we assume a mean vector density (number of mosquitoes per CDC light traps) of 3.3, a standard deviation (SD) of 3.3, and CV of 0.192. For 80% power to detect a 50% decrease in mean mosquito densities at 5% type 1 error level, we need to capture mosquitoes from five houses per cluster.

### Recruitment {15}

Thirty-eight eligible children in each cluster will be randomly recruited into our cohort. Recruitment will be limited to children aged 12 or younger, to avoid children aging out during the 18-month monitoring period. Study team staff will obtain informed consent from the parents or caregivers of the children before enrolling the children in the cohort. For the cross-sectional survey at each time point, we will randomly select from each cluster 50 individuals of all age groups. To guarantee the representativeness for all age groups, the selection will be done with following age category stratifications; 0-4, 5-9, 10-14, 15-19, and 20 and above.

### Assignment of interventions: allocation Sequence generation {16a}

Random numbers will be generated using the sample function in R software.

### Concealment mechanism {16b}

The individual, household, and villages (clusters) are all given unique IDs at the beginning of the baseline. Any following steps handle only these anonymized IDs.

### Implementation {16c}

After the baseline survey, covariate-constrained randomization will be used to allocate the 44 clusters across the two study arms. The following factors will be constrained: baseline malaria infection prevalence by RDT in children aged 0.5–14 years, LLIN usage, malaria vaccine coverage, socioeconomic status (SES), population size, the proportion of eligible houses for the ceiling net installation, and vector densities. An independent statistician will perform the randomization. Local study assistants will perform participant enrollment.

### Assignment of interventions: Blinding Who will be blinded {17a}

Due to the visibility of the Olyset®Plus ceiling net, neither the trial participants nor the members of the study team who take part in field activities can be blinded. However, laboratory- and office-based personnel (e.g., microscopists, laboratory technicians, and data analysts) will be blinded to the identity and intervention status of the trial participants since all biological specimens will be identified by a unique numeric study identifier, and personal information will be removed before analyses.

### Procedure for unblinding if needed {17b}

This is an open-label trial, and only the data measurers are blinded. Therefore, there is no circumstance that they need to be unblinded.

## Data collection and management

### Plans for assessment and collection of outcomes {18a}

#### Baseline survey

A baseline survey targeting 50 enumeration areas (comprising one or two villages) in the study area will be conducted shortly before the ceiling net intervention to obtain data to assure the balance of the cluster allocation and obtain basic demographic data. The baseline survey includes a questionnaire for all households, mRDT testing of all children aged 6 months to 14 years, and an entomological survey of randomly sampled households. We modified the questionnaire used in the 2020 Kenya Malaria Indicator Survey (KMIS) mainly to quantify the SES of each household and bed net usage. In addition, we add questions to quantify the favorability of ceiling nets before the intervention.

#### Cohort monitoring

Incidence of clinical malaria in the prospective cohort will be estimated by both active and passive case detections. For active case detection, we will conduct home visits every two weeks. Axillary temperature will be measured using a digital thermometer. Participants with fever (>37.5°C) or other malaria-related symptoms listed in the Kenya National Malaria Treatment Guideline at the time of home visit or within the previous 48 hours will be tested for malaria by RDT. History of travel, confirmed malaria episode, and visit to local health facilities since the previous visit is recorded. *Plasmodium* infection status will be determined by RDT and PCR from all cohort participants during every other biweekly home visit i.e. every four weeks. For passive case detection, we ask all cohort participants to visit designated health facilities in case they suspect malaria between home visits. The designated facilities are asked to record all malaria tests performed regardless of their results together with the cohort ID. The cost of RDT and anti-malarial treatment will be covered by the research team to encourage cohort participants to use only designated facilities.

#### Cross-sectional malariometric surveys

Malaria prevalence in children and adults will be estimated using cross-sectional malariometric surveys in communities. These surveys will be conducted at 6, 12, and 18 months post-installation. Community surveys will be conducted by house visits.

*Plasmodium* infection status will be determined using three methods: RDT, microscopy, and PCR. A finger-prick blood sample will be collected for on-site diagnosis using the Bioline Malaria Ag P.f/Pan RDT (Abbott Diagnostics Korea Inc., Republic of Korea). Survey participants with positive test results will be provided with a treatment course of artemether-lumefantrine with dosing instructions in accordance with guidelines from the Ministry of Health in Kenya after checking their recent treatment history. Blood smears will be prepared on site and transported to the main laboratory in Homa Bay where thin smears are fixed with methanol and all smears are stained with 3% Giemsa solution for 30 minutes, then examined by experienced microscopists. Two blood samples (70 µl each) will be collected with a 75-mm heparinized micro-hematocrit capillary tube (Thermo Fisher Scientific, MA, USA) and spotted on Whatman ET31 Chr filter paper (Whatman International. Maidstone, UK). The blood samples will be allowed to dry at ambient temperature and stored in individual zipped plastic bags at −20□. The dried blood spots will be used for the determination of malaria status by PCR [17].

#### Entomological surveys

Indoor resting mosquitoes will be collected from five sentinel houses within each cluster using the CDC light trap method. Samples will be preserved in 96% ethanol and placed in a cool box with ice. Specimens will be examined for sex determination by microscopy and species identification by microscopy and PCR. Indoor resting mosquitoes will be collected at baseline, 6, 12 and 18 months post ceiling net installation.

#### Social aspects

We will conduct an exploratory sequential research design using integrated mixed methods (qualitative and quantitative). Qualitative assessment of community perceptions on the Olyset®Plus ceiling nets, community facilitators, and concerns of Olyset®Plus ceiling net use will be implemented, followed by quantitative assessments every 6 months and routine monitoring to evaluate durability and appropriateness of fit of Olyset®Plus ceiling nets using observation checklists. At the end of the study, other qualitative case studies, such as focus group discussions and key informant interviews, will be conducted to document success stories and inform the sustainability and scalability of the intervention.

#### Cost-effectiveness analysis

Incremental financial and economic cost data of Olyset®Plus ceiling net will be collected alongside the intervention. In cases where resources, such as staff, are shared among multiple elements, the allocation of costs will be carried out using an appropriate proxy. Costs related to research activities will be excluded from this allocation. Financial costs will be derived from project expenditure records, while economic costs, which encompass financial expenditures and donated resources, will be identified through project records and social aspects activities. The value of donated resources will be credited based on prevailing market rates. Furthermore, capital costs will be annualized over their useful life for financial costing and annualized at a discount rate of 3% for economic costing.

### Plans to promote participant retention and complete follow-up {18b}

All surveys planned for the epidemiological and entomological domains will be conducted by house visits. CHPs will make an appointment with eligible participants before each visit to confirm the participants’ available date and time. Small remunerations will be provided to survey participants to compensate for their time. CHPs will receive detailed instructions and participatory training for all field procedures and will be actively supervised by the research team throughout the duration of the study. Feedback will be regularly sought from CHPs regarding any issues raised by study participants, and discussions will be held to resolve issues from the field.

### Data management {19}

All data from the baseline, cohort, and cross-sectional surveys will be captured using the Research Electronic Data Capture (REDCap) software on electronic tablets. Data will be uploaded daily to a highly secure server hosted by Mount Kenya University (MKU). All data from the quantitative surveys will also be stored securely and backed up regularly to prevent data loss. Data access and management of databases will be limited to authorized study investigators and collaborators. After validation of data uploaded to the MKU server, data stored locally on the tablet computers will be permanently deleted to minimize unauthorized access.

### Confidentiality {27}

To maintain confidentiality, each participant in cross-sectional surveys, the longitudinal cohort, and the quantitative surveys is assigned a unique identifier. The data collected will be labelled using the unique identifier and stored separately from the key linking personal information (name, date of birth, GPS of each household, and phone number). The data will be kept on a secure server that is only accessible to the research staff. Publications will contain only aggregated data, and no personal information will be included.

### Plans for collection, laboratory evaluation and storage of biological specimens for genetic or molecular analysis in this trial/future use {33}

Anonymized blood samples from study participants will be stored and analyzed for *Plasmodium* infections by microscopy and PCR in our laboratory in Homa Bay. Microscopic examinations of adult mosquito specimens will be conducted in our field laboratory in Mbita, while PCR analyses will be conducted in our laboratories in Homa Bay and MKU. Laboratory data outputs will be entered in Microsoft Excel and imported into the database. No human genetic analysis is planned for this study. However, remaining biological materials will be stored indefinitely for future studies unless the participants opt out during the informed consent process. Participants are provided with contact information of the research team and can remove themselves from this study or any future studies at any time without penalty ore prejudice.

## Statistical methods

### Statistical methods for primary and secondary outcomes {20a}

We will follow the CONSORT guidelines extended for CRCT for statistical analysis and result reporting. The intention-to-treat (ITT) analysis is the primary analysis approach for both the primary and secondary objectives for the epidemiological and entomological studies. The per-protocol (PP) analysis is included as a supplementary analysis for the primary and secondary objectives for the epidemiological and entomological studies. Detailed methodologies for the epidemiological part are described in the supplementary file of statistical analysis plan.

#### Clinical malaria incidence

We will determine the protective efficacy of Olyset®Plus ceiling nets against malaria case incidence by comparing clinical malaria incidence rates between arms. We will use mixed effects negative binomial regression accounting for within-cluster correlation of outcomes. Possible confounding factors such as age, sex, bed net usage, house structure, malaria vaccination history, and SES will be adjusted as well as covariates used in the covariate-constrained randomization. In addition, because we will not set a buffer zone, the distance to the nearest household in the other arm will be adjusted in the following analysis to reduce the contamination between two arms. The variable was selected from the previous study [18].

#### Prevalence of malaria infection

The secondary outcome, the prevalence of malaria infection by PCR and microscopy measured at 6, 12, and 18 months after the ceiling net installation will be analyzed using mixed effects logistic regression adjusting for the above-mentioned confounding factors.

#### Time to first malaria infection

A Cox proportional hazards model and other survival models will be used to compare time to first malaria infection between arms adjusting for the above-mentioned confounding factors. In addition, we will account for the within-cluster correlation of responses.

#### Exploratory analysis for spillover effects

Evidence for positive spillover effects of the ceiling net on malaria infection prevalence of all age group will be assessed by comparing individuals with no intervention conditioning 1) the distance to ceiling net installed household, and 2) the coverage of surrounding households with ceiling net within 400 m. The distance of 400 m was chosen as the spillover effect appears to attenuate at this distance based on previous reports [19].

#### Entomology

Differences in vector density and EIR between arms will be evaluated by random effects negative binomial regression taking into account the intracluster correlation.

#### Social aspects

We will employ the Framework for Reporting Adaptations and Modifications to Evidence-based Implementation Strategies (FRAME-IS) [20] to document the implementation processes of the ceiling nets, and the Evidence integration triangle framework[21] to align the evidence generated to policy and vector control strategies from the health systems aspect. The theoretical framework in qualitative research will be grounded theory[22]. Data from ethnographic, focus group discussions, key informant interview will be summarized using content thematic analysis. Pre- and post-intervention acceptability to install Olyset®Plus ceiling net intervention will be compared to actual consent using logistic regressions.

#### Cost-effectiveness

The economic and financial costs associated with the Olyset®Plus ceiling net intervention will be presented in total and disaggregated forms, highlighting the relative contribution of each program element to the overall program costs. To facilitate comparisons with other malaria vector control interventions, the costs will be converted into cost per household and per person receiving the intervention annually. Various program scenarios, such as different scales and durations, will be presented to estimate operational implementation costs. Compared to the control group, we will utilize the number of malaria cases averted in the Olyset®Plus ceiling net arm to calculate the DALYs averted using standard methods.

### Interim analyses {21b}

No interim analysis is planned because neither the insecticide permethrin nor the synergist PBO as formulated in Olyset®Plus LLINs are known to pose significant health and safety risks [9,23].

### Methods for additional analyses (e.g. subgroup analyses) {20b}

We will perform the same analysis for three age subgroups (≤ 59 months old; 5 years old to 14 years old; 15 years old or older) to examine if the effects of Olyset®Plus ceiling net differ by age groups. In addition, we will perform other machine learning based approach such as causal forest and super learner to estimate the conditional average treatment effect.

### Methods in analysis to handle protocol non-adherence and any statistical methods to handle missing data {20c}

In the cohort, non-adherence to the intervention can be identified by bi-weekly interviews. Participants who regularly sleep outside their homes will be removed from the analyses. The extent and patterns of missing data will be assessed once all data collection has been completed. If necessary, we will apply simple hot-deck imputation methods if the missing fraction for the covariate is <5% or appropriate multiple imputation approaches if the missing fraction for a covariable are ≥5%. If a non-ignorable portion of the subjects have missing values on a covariate (due to missing at random or missing completely at random), that covariate may be excluded in the model.

### Plans to give access to the full protocol, participant level-data and statistical code {31c}

This manuscript is the full protocol. The corresponding author will make the de-identified datasets or any future statistical code available upon reasonable request.

### Oversight and monitoring

#### Composition of the coordinating centre and trial steering committee {5d}

The sampling team, composed of CHPs and laboratory technicians, set up a day-to-day communication group and exchanged their experiences. A local management team of study investigators from Kenya and Japan also joined this, leading and advising the activities and monitoring the sample and data integrity. A monthly meeting will be held by the steering committee composed of all key researchers from Kenya and Japan, including the principal investigator (PI) and co-PI, which aim to monitor the progress of the trial.

### Composition of the data monitoring committee, its role and reporting structure {21a}

Because this intervention is considered to be of a low-risk nature, this study does not have a data monitoring committee. For additional credibility about study quality, the researchers will consult a third statistician, if necessary.

### Adverse event reporting and harms {22}

All unanticipated problems will be reported to the research team and Homa Bay County Ministry of Health (MOH) through CHPs. Medical officers from Homa Bay County will assess the relatedness of the reported events to the study and report to the research team, including the PI. In the event of a study-related serious adverse event, the study team will convene a meeting immediately with the MOH and Homa Bay County Teaching and Referral Hospital representatives to review the case and take necessary action. Also, the ceiling net is made of the same materials and chemicals as LLIN already on the market, and is therefore not expected to have significant environmental impact.

### Frequency and plans for auditing trial conduct {23}

A monthly meeting will be held during the follow-up period to ensure that all surveys and investigations are conducted according to the study protocol. The study is required to submit annual reports and renewal to ethical review boards of Osaka Metropolitan University, Japan, and Mount Kenya University, Kenya.

### Plans for communicating important protocol amendments to relevant parties (e.g. trial participants, ethical committees) {25}

Decisions on important trial amendments must be made through a formal procedure and will be approved by institutional review boards (IRB) at Mount Kenya University and Osaka Metropolitan University. The protocol in the clinical trials registry will also be updated accordingly.

### Dissemination plans {31a}

Study results will be shared with the study participants and communities, the Homa Bay County Government and the Kenya National Malaria Control Programme. Results will also be disseminated through publications, conferences and workshops to help the development of novel malaria control strategies in other malaria-endemic countries. Suggestions from the participants will also help shape the future improvement of the intervention.

## Discussion

Global malaria progress has flatlined in recent years: targets of reductions in malaria morbidity and mortality and required funding by 2030 are all off track as of 2023[6]. In addition, *P. falciparum* with partial artemisinin resistance, which has been a problem in the Great Mekong Subregion (GMS) for more than a decade [24] [2–6]. Novel interventions that are cost-effective and widely accepted by local communities are urgently needed to contain the spread of artemisinin-resistant *P. falciparum* in sub-Saharan Africa.

Early results from our cluster randomized controlled trial of Olyset®Plus ceiling nets on Mfangano Island in Lake Victoria, Kenya suggest that ceiling nets can reduce *Plasmodium* prevalence and are positively received by the local communities. Nevertheless, there are regional differences in housing design, vector abundance and composition, and availability of malaria control interventions. As such the feasibility and acceptability of the ceiling net intervention are likely to depend on local eco-epidemiological context [25]. Furthermore, one of the secondary objectives in this study is to measure the spillover effects, i.e. how much a household that does not have a ceiling net benefits from living near a house with a ceiling net. This enables a broader understanding of the impact of the ceiling nets at the community level.

This trial has several limitations. First, although the study is designed as a cluster-randomized controlled trial, contamination between intervention and control clusters cannot be excluded, as buffer zones between intervention and control clusters cannot be created due to geographic proximity of houses and villages in the ward. A recent study, however, has shown that the spillover effect of interventions on malaria can extend to 3 km [26], so buffer zones of a few hundred meters, as set out in many studies, may not be sufficient. We will try to eliminate such contamination effects by integrating spatial data into our statistical model. Second, because of the visible nature of the ceiling net, we cannot exclude open-label and observer biases. It is conceivable that participants receiving ceiling nets may reduce their usage of conventional LLIN, as both interventions are made of the same materials and may be perceived to protect against malaria in the same manner. We aim to reduce such bias as much as possible through repeated reminders by CHPs that ceiling nets serve as an addition to and not a replacement of conventional LLINs. We will conduct surveys and in-depth interviews to elicit participants’ perceptions of the ceiling net, which can guide future messaging and implementation. To reduce observer bias, laboratory investigators and data analysts will be blinded. Third, in the study area, pyrethroid+PBO-incorporated LLINs were distributed in 2023. It may reduce the effect of our ceiling net intervention because pyrethroid+PBO-incorporated LLINs are more effective than non-PBO-incorporated LLINs by targeting both *Anopheles* vectors with and without metabolic resistance to pyrethroids. Pyrethroid+PBO incorporated LLINs received a conditional endorsement from the World Health Organization (WHO) in 2017, and approximately half of the LLINs distributed in sub-Saharan Africa in 2022 were of this type [2]. Given the abundance of PBO-incorporated LLINs in the region, it is important to assess the effectiveness of the Olyset®Plus ceiling net as an addition to these LLINs to inform policy recommendations. Recently LLINs combining two different classes of insecticides have been shown to be superior to pyrethroid-based LLINs [27]. When these new LLINs become widely available, the effectiveness of pyrethroid-PBO ceiling nets needs to be re-investigated.

## Trial status

The Baseline survey was started on April 8, 2024. The recruitment of the intervention participants will be in June 2024. The current protocol is version 5.0 as of April 18.

## Supporting information

Study Analysis Plan

## Data Availability

The study regimes, consent forms, assent forms, and study-related materials are accessible from the corresponding author. The final trial dataset will be available to all investigators. The corresponding author will make the de-identified datasets and source codes for all analysis available upon reasonable request.

## Abbreviations

ACT: artemisinin-based combination therapy
CHP: community health promoter
CRCT: cluster-randomized controlled trial
CV: coefficient of variation
*kdr*: knockdown resistance
KHIS: Kenya Health Information System
ITN: insecticide treated nets
IRS: indoor residual spraying
LLIN: long-lasting insecticidal nets
MKU: Mount Kenya University
PBO: piperonyl butoxide
RDT: rapid diagnosis tests

## Declarations Acknowledgements

We would like to express our sincere gratitude to this study’s participants, field, and laboratory staff. In addition, we acknowledge the collaboration and support of health offices in Homa Bay County, Kenya.

## Authors’ contributions {31b}

AK and JG are co-principal investigators. YKK, WK, and AK developed the original concept. All authors discussed and contributed to the study protocol. YKK, WK, PO, BM, TO, CWC, JK, SM, and MK drafted the manuscript. YKK, WK, CWC, MK, GO, and JG contributed to the revisions of the draft of the manuscript. DY participated as a senior statistician. YKK and MK drafted the statistical analysis plan (SAP) and WK, CWC, and DY revised. The authors read and approved the final manuscript and SAP.

## Patient and public involvement

Although the study design was developed through discussions among the researchers, consultations with the local population were conducted prior to initiating the baseline survey, and their input was incorporated into the study. Community involvement will also be ongoing during the implementation of interventions and research activities.

## Ethics approval and consent to participate {24}

Ethics approval was received from Mount Kenya University Institutional Scientific Ethics Review Committee (MKU-ISERC) and is under the review from the Ethics Committee in Osaka Metropolitan University.

Written informed consents will be sought from study participants before the baseline survey, installation of ceiling nets, each cross-sectional survey, and the start of prospective cohort surveys. Participants have the right to withdraw from the study at any time, and the option to withhold previously collected samples from any future analyses and studies.

The samples collected in this study may potentially be used for other research purposes. This is clearly stated in the informed consent form. In such cases, we will obtain the necessary ethical approval and provide participants with the chance to opt-out from this. All experiments will be carried out in adherence to WHO requirements and the Declaration of Helsinki.

## Consent for publication {32}

We will not present identifying images or other personal or clinical details of participants. The participant information materials and informed consent form are available from the corresponding author on request.

