## Supplementary material for "Evaluation of the protective efficacy of Olyset®Plus ceiling nets for reduction of malaria incidence in children in Homa Bay County, Kenya: a cluster-randomized controlled study protocol": Study Analysis Plan

Evaluation of the protective efficacy of Olyset®Plus ceiling nets for reduction of malaria incidence children in Homa Bay County, Kenya: statistical analysis plan for clinical and epidemiological outcomes

**SAP version**

Version 4.0

Apr 9, 2024 prepared by Yura K Ko

**SAP revisions**

| Version | Date | Summary of Changes |
| --- | --- | --- |
| 1.0 | Jan 23, 2024 | First draft |
| 2.0 | Feb 5, 2024 | Added more details based on collaborators’ feedback |
| 3.0 | Mar 4, 2024 | Revised by the senior statistician (Dr. Daisuke Yoneoka) |
| 4.0 | April 9, 2024 | Revised based on collaborators’ feefback |

### Introduction

#### Objectives

*Primary objective*:

- To determine the protective efficacy of Olyset®Plus ceiling nets in reducing malaria case incidence in children 6 months-14 years for 18 months post-intervention

*Secondary objectives*:

1. To determine the protective efficacy of Olyset®Plus ceiling nets in reducing malaria infection prevalence in all age groups at 6-, 12-, and 18-months post-intervention
2. To determine the protective efficacy of Olyset®Plus ceiling nets against the time to first malaria infection for 18 months post-intervention
3. To determine the spillover effects of Olyset®Plus ceiling nets in reducing malaria infection prevalence in all age groups at 6-, 12-, and 18-months post-intervention
4. To determine the protective efficacy of Olyset®Plus ceiling net in reducing *Plasmodium* infection incidence in children 6 months to 14 years old over 18 months post-intervention.

### Study Methods

#### Trial design

The study is a cluster-randomized controlled trial (CRCT) with 44 clusters evenly divided between the intervention and the control arms. Each cluster will be one or two villages consisting of at least 50 households. A baseline survey will be conducted to determine pre-intervention *Plasmodium* prevalence and to collect demographic and socioeconomic data for covariate-constrained randomization of clusters. The baseline survey will be conducted approximately one month before randomization. The post-intervention follow-up period will be 18 months. Thirty-eight children aged 6 months to 14 years from each cluster will be recruited and followed for 18 months as a cohort to determine the protective efficacy of the Olyset®Plus ceiling net on clinical malaria incidence (primary objective), time to first *Plasmodium* infection, and *Plasmodium* infection incidence (secondary objectives). Cross-sectional surveys will be conducted at 6, 12, and 18 months post-intervention targeting 50 individuals of all ages from each cluster to determine the overall *Plasmodium* prevalence and to estimate the spillover effect (secondary objectives).

#### Randomization

After the baseline survey, covariate-constrained randomization will be used to allocate the 44 clusters across the two study arms. Covariate-constrained allocation ensures that the arms are balanced overall by excluding allocations where predetermined factors are not balanced within set margins. The following factors will be constrained: baseline malaria infection prevalence by RDT in children aged 0.5–14 years, LLIN usage, malaria vaccine coverage, socioeconomic status (SES), population size, the proportion of eligible houses for the ceiling net installation, and vector densities.

For the proportions of interest, we require the average difference between the arms of no more than 10% for the means of interest, and we require the difference in means to be no more than a quarter of the standard deviation of the variable among individuals in the population. After enumerating the allocations that fulfil the criteria, we may relax or tighten up the balance criteria when the allocated number is too small or very large. One allocation will be selected randomly among all possible allocations meeting the balancing constraints. Data on any additional potentially confounding ecological factors not included in the covariate-constrained randomization will be collected and adjusted for in the analysis. An independent statistician will perform the randomization.

#### Sample size

The sample size was calculated using the method of Hayes and Moulton^1^. All the sample sizes will be recalculated based on the baseline data, which will be collected before the ceiling net installation.

The following calculations were based on the historical data collected from the same Lake Victoria region in Kenya with a clinical malaria incidence rate of 0.5 per person-year for children under 14 years old by RDT (personal communication), 40% parasite prevalence for all age groups by PCR, and a between-cluster coefficient of variation (CV) of incidence rate = 0.24 in both arms. In the study site, RTS, S vaccination began in 2019, with an additional mass distribution of pyrethroid-PBO LLINs at the end of 2023. Therefore, the intervention effect is expected to be smaller than in previous studies and was conservatively assumed to be 25%. Assuming 38 individuals per cluster to be followed for up to 18 months with 20 % loss-to-follow up rate, we require 22 clusters per arm to achieve 80% power to detect a significant incidence rate ratio of 0.75 (25% protective efficacy) at a two-sided type 1 error of 5%. With 44 clusters and 50 individuals per cluster, for the outcome of *Plasmodium* prevalence by PCR, we would achieve 80% power to detect a 23.5 % relative reduction. We do not specify the sample size for identifying spillover effects because spillovers tend to have smaller effect sizes relative to total or overall effects, so typically larger sample sizes are required to detect them. Although our study may be underpowered to detect spillovers, we will report the results as an exploratory analysis.

#### Framework

Our null hypothesis for the primary outcome is that Olyset®Plus ceiling nets + standard malaria treatment and prevention measures do not reduce the clinical malaria incidence compared to standard malaria treatment and prevention measures in children 6 months to 14 years at 18 months post-intervention.

#### Statistical interim analyses and stopping guidance

Neither the ceiling nets nor synergist PBO are known to pose significant health and safety risks. This has also been demonstrated in our previous CRT on Mfangano Island^2^. Therefore, no interim analysis is planned.

#### Timing of final analysis

We will conduct the final analysis after 18 months of follow-up. Results will immediately be submitted for publication in peer-reviewed journals.

#### Timing of outcome assessments

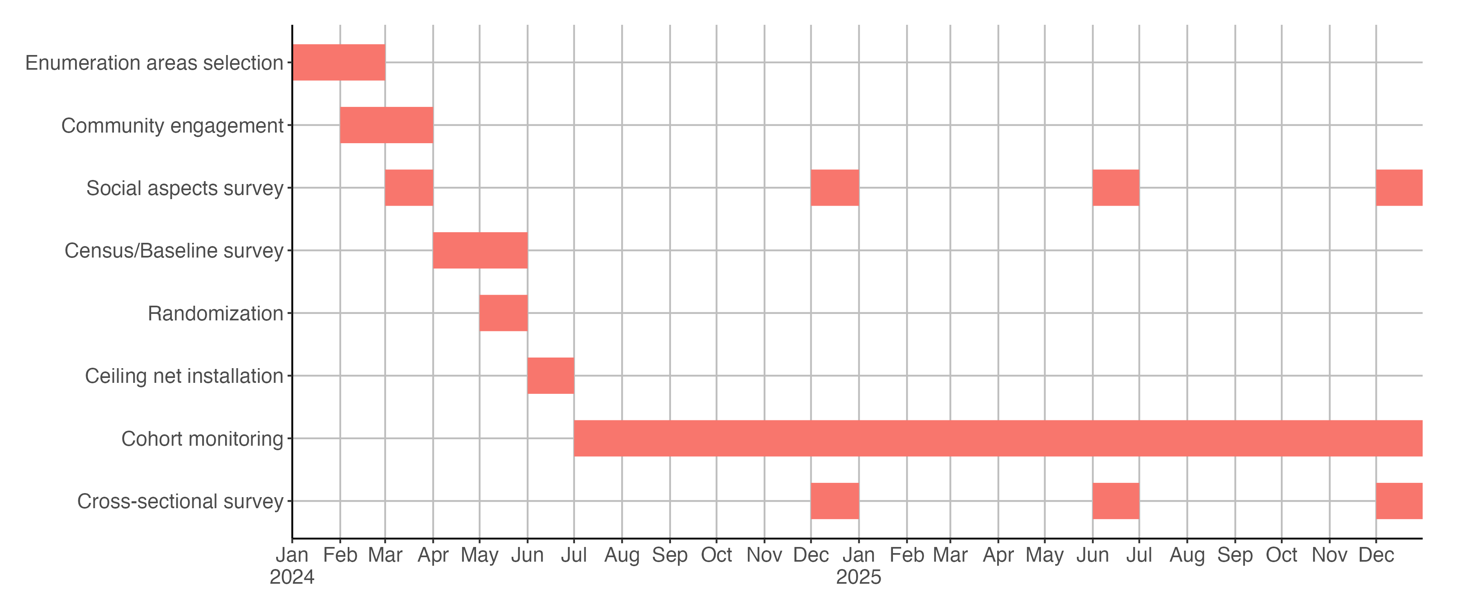

Figure1: Timetable of trial activities

### Statistical Principles

#### Confidence intervals and P values

We will use a two-sided significance level of α = 0.05 for hypothesis testing. All statistical tests will be conducted at this predetermined level of significance unless otherwise specified. For each estimated parameter, 95% confidence intervals will be calculated and reported.

#### Adherence and protocol deviations

Since the intervention (ceiling net) will be installed in trial participants’ houses, participants not sleeping in their own houses will not benefit from the intervention. In the cohort, non-adherence to the intervention can be inferred from travel history during bi-weekly interviews. Therefore, participants who regularly sleep outside their homes will be removed from the analyses. In addition, if a cohort participant is absent for three consecutive visits or for more than half of all visits, the child will be excluded from the analysis. Other protocol deviations will be carefully documented and categorized during the study.

#### Analysis populations

The intention to treat (ITT) analysis is the primary analysis approach for both the primary and secondary objectives. The per-protocol (PP) analysis is included as a supplementary analysis for the primary and secondary objectives.

### Trial population

#### Screening data

Screening data will be collected during the cohort enrollment and each cross-sectional survey to assess the eligibility of potential participants. This information will include demographic characteristics such as age, sex, SES, and other relevant parameters outlined in the study protocol.

#### Eligibility

The inclusion criteria for the installation of the Olyset®Plus ceiling net are (1) residential structures housing at least one permanent resident aged 18 years or older in the household, (2) informed consent provided by at least one adult in the household and (3) applicable house structure for the ceiling net in terms of size of the structure, presence of eave, ceiling board, and vertical beams, and material of the top part of the wall. The applicability of the ceiling net installation will be assessed by experienced field staff. The exclusion criteria are (1) vacant dwelling structure (confirmed by at least two visits by CHPs), (2) dwelling structure to be vacated or destroyed within the study period and (3) not applicable house structure for the ceiling net installation.

The inclusion criteria for prospective cohorts of children aged 6 months to 14 years old are (1) living in the study area at the time of Olyset®Plus ceiling net installation, (2) having no plan to leave or stay outside the study area for an extended period (longer than 1 month) over the 18-month follow-up period, and (3) informed consent provided by the participant’s parent or guardian. The exclusion criterion is having severe chronic illnesses.

The inclusion criteria for cross-sectional malaria surveys for all age groups are (1) living in the study area during the study period, and (2) informed consent being provided by the parent or legal guardian before each survey. The exclusion criterion is having severe chronic illnesses.

#### Recruitment

For the baseline survey, we will randomly choose 50 enumeration areas (comprising one or two villages) in Kanyamwa Kologi Ward, Ndhiwa Sub-County. The survey includes a questionnaire for all households, mRDT testing of all children aged 6 months to 14 years. We will not create buffer zones to minimize contamination since a buffer zone of 400–600 m from the boundary will greatly reduce the number of houses in the core area available for analysis in many clusters. Thirty-eight eligible children will be randomly recruited into our cohort in each cluster. Recruitment will be limited to children aged 12 or younger, to prevent children from aging out during the 18 months monitoring period. For the cross-sectional survey at each time point, we will randomly select 50 individuals of all age groups from each cluster. To reflect the age structure of the populations, the selection will be done with age category stratifications.

#### Withdrawal/follow-up

Those who migrate between the arms or emigrate from the study areas, or those who dismount the ceiling net from their house structure will be dropped from the intervention. For the cohort, those who are absent for three consecutive visits or for more than half of all visits will be excluded from the analysis as lost to follow-up. A summary of study participant selection is shown in Figure 2.

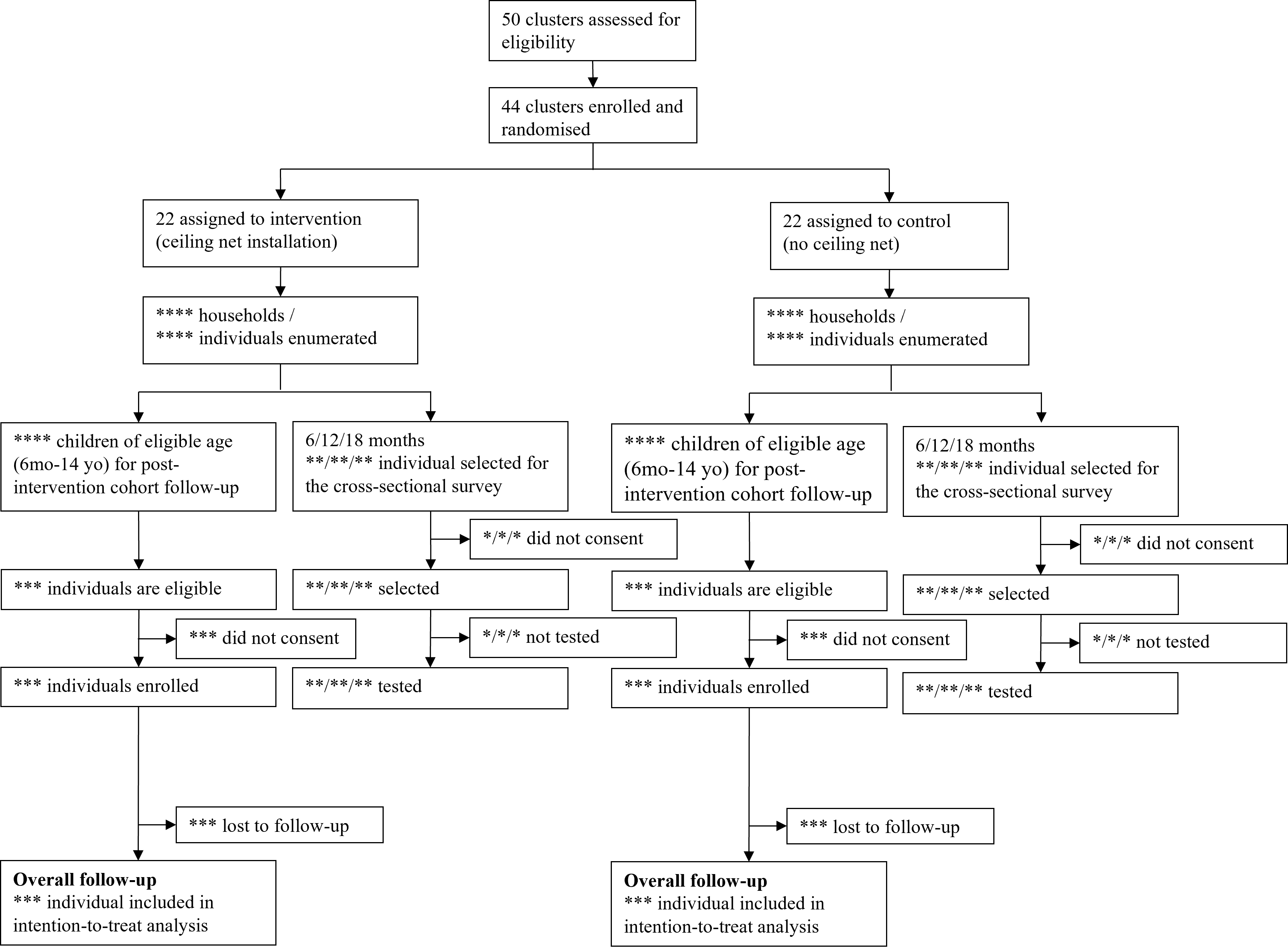

Figure 2: A schematic flow diagram of cluster allocation and study participant selection.

#### Baseline characteristics

We will report a list of baseline characteristics of the intervention and control arms. The list will include population, mean number of people per household, median age of population, number of selected children for the cohort monitoring, number of selected individuals for each cross-sectional survey, malaria infection prevalence by RDT in children aged 0.5–14 years, LLIN usage, malaria vaccine coverage among children, SES, the proportion of suitable houses for the ceiling net installation, the mean indoor vectors per household per night.

### Analysis

#### Endpoints

*Primary endpoint*:

- Overall malaria case incidence in children 6 months-14 years among intervention and control clusters during the 18-month follow-up period.

*Secondary endpoints*:

1. Malaria infection prevalence at 6, 12, and 18 months post-intervention in all age groups among intervention and control clusters.
2. Time to first malaria infections in children 6 months-14 years among intervention and control clusters during the 18-month follow-up period.

#### Definition of malaria case incidence

Incidence of clinical malaria in the prospective cohort will be estimated by both active and passive case detections. For active case detection, we will visit the home of cohort participants every two weeks. At each biweekly visit, axillary temperature will be taken from each cohort participant. If the participant has fever (>37.5°C) or any malaria-related symptoms during or within 48 hours of the home visit, the child will be tested by mRDT. A clinical case is defined as positive mRDT accompanied by fever and/or any malaria-related symptoms and will be treated with artemether-lumefantrine (artemisinin-based combination therapy [ACT]). For passive case detection, we ask all cohort participants to visit designated health facilities in case they suspect malaria between home visits.

If two consecutive RDTs are positive, there are two patterns: active case detection or passive case detection for the detection of the second positive. If the second positive RDT is detected by active case detection, we will refer the child to a health facility and regard it as a new malaria infection if subsequent microscopy or PCR confirms parasites after 15 days or more passed from the first RDT test. If not, it is considered a carryover from the previous infection. For the passive case detection of a second RDT positive, we will regard it as a new malaria infection if more than 14 days have passed since the first RDT test.

For the overall incidence rate calculation, the time at the risk will be adjusted by subtracting the 14 days of “protection period” of ACT. If a participant misses a particular visit or a second positive is considered a carryover from the previous infection, the period is not included in the at-risk period.

If the cohort children visit health care facility and get tested for malaria between each visit, both positive and negative results will be utilized for our analysis. Specifically, the 14 days prior to the test result will be incorporated into the denominator of the incidence calculation as the at-risk period. Specific patterns for incidence rate calculation are shown in Figure 3.

If a participant is absent for three consecutive visits or for more than half of all visits, the child will be excluded from the analysis.

#### Definition of malaria infection incidence

In addition, we will test all cohort children by RDT and PCR every month. For the secondary outcome of time to first malaria infection and infection incidence, because our focus is only on infection, not symptomatic infection. We therefore use passive case detection differently to the above. If the cohort children visit health care facility because of any symptom and get negative RDT results for malaria, this data point is removed as it does not provide any additional information. If the RDT results in the facility is positive, the passive positive is assigned to either the active visit immediately before or after the passive case detection, whichever is closer in time to the passive case detection.

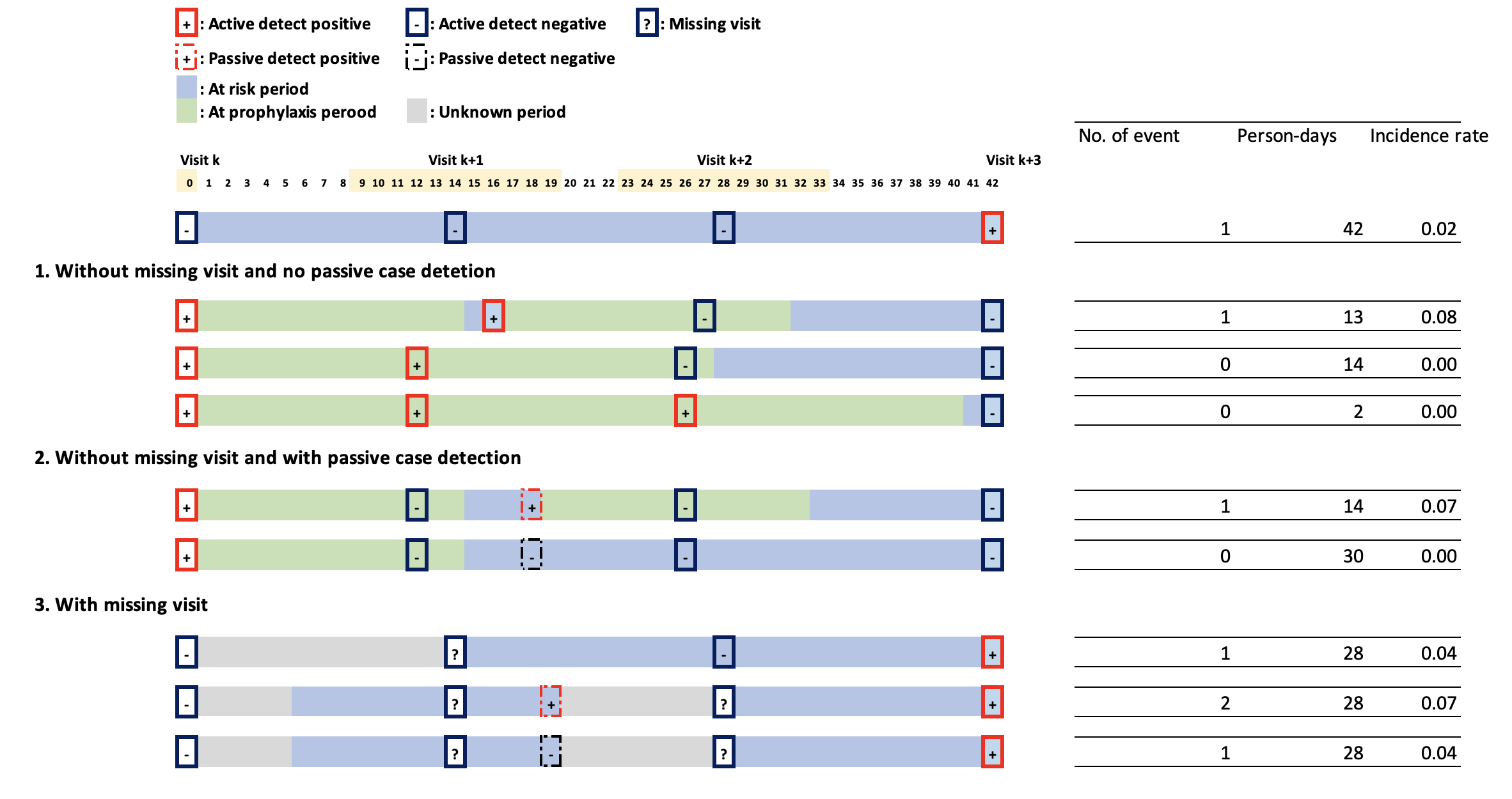

Figure 3: Specific patterns for incidence rate calculation.

#### Analysis methods

We will follow the CONSORT guidelines extended for CRT^3^ for the statistical analysis and results reporting.

*Clinical malaria incidence*

We will determine the protective efficacy of Olyset®Plus ceiling nets against malaria case incidence by comparing clinical malaria incidence rates between arms. We will use mixed effects negative binomial regression accounting for within-cluster correlation of responses by following equations:

$\mu_{k,i}=exp(\beta_{0}+\boldsymbol{x}_{k,i}^{T}\boldsymbol{\beta}+z_{k})$,

$z_{k} \sim N(0,\sigma^{2})$,

where $\mu_{k,i}$ is the mean incidence rate of individual $i$ in cluster $k$, $\boldsymbol{x}_{k,i}$ is the covariate vector including individual and cluster level data, $z_{k}$ is the Gaussian-type random effect at the cluster level. The protective efficacy will be estimated by $\left( 1-\exp\left( \hat{\beta} \right) \right)\times100\%$, where $\hat{\beta}$ is the estimated regression coefficient of the treatment. Possible confounding factors such as age, sex, bed net usage, house structure, and SES will be adjusted as well as the covariates used in the covariate-constrained randomization. In addition, because we will not set a buffer zone, the distance to the nearest household in the other arm will be adjusted as a covariate to reduce the contamination between two arms. The variable was selected from the previous study^4^.

*Prevalence of malaria infection*

The secondary outcome, prevalence of malaria infection by PCR and microscopy measured at 6-, 12-, and 18-months after the ceiling net installation will be analyzed using mixed effects logistic regression adjusting for the above-mentioned confounding factors.

*Time to first malaria infection*

A Cox proportional hazards model and other survival models will be used to compare time to first malaria infection between arms adjusting for the above-mentioned confounding factors. In addition, we will account within-cluster correlation of responses.

*Exploratory analysis for spillover effects*

Evidence for positive spillover effects of the ceiling net on malaria infection prevalence of all age group will be assessed by comparing individuals with no intervention conditioning 1) the distance to ceiling net installed household, and 2) the coverage of surrounding households with ceiling net within 400 m (Figure 4). The distance of 400 m was chosen as the spillover effect appears to attenuate at this distance based on previous reports^5^. As there may be a bias that households without a ceiling net in the intervention cluster have different characteristics (e.g. preventive behaviour against malaria), we will include only control clusters for the spillover analysis to ensure comparability.

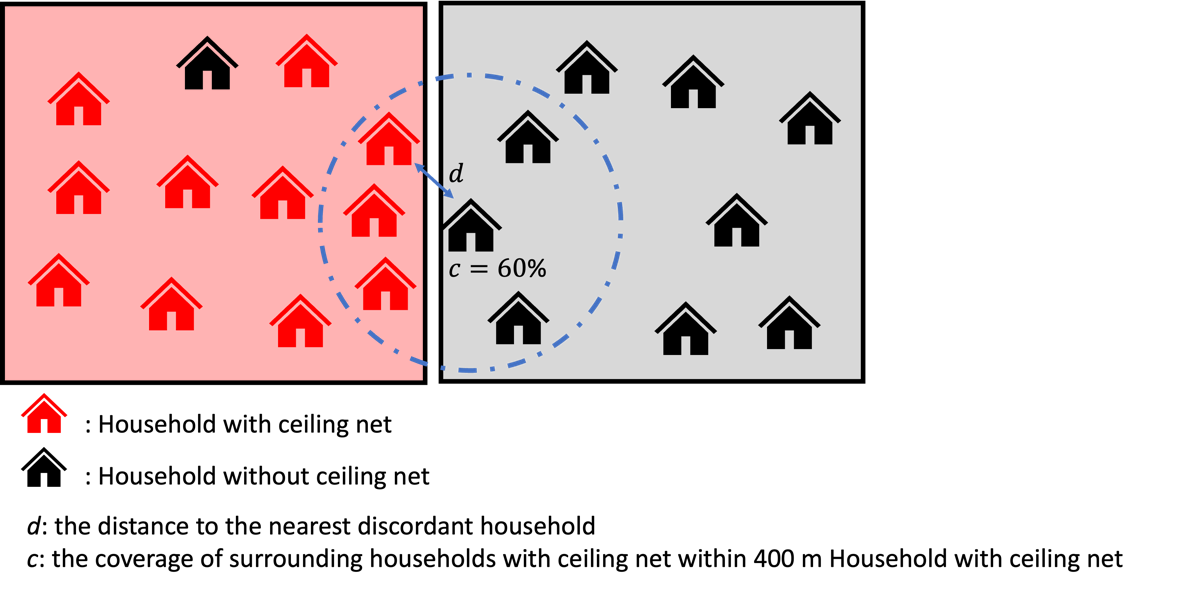

Figure 4: The distance to the nearest discordant household and the coverage of treatment.

#### Missing data

We will make substantial effort to avoid having missing values on outcome (malaria infection status and visit dates) by encouraging individual participants and CHPs repeatedly. When missing values occur for an outcome for reasons not related to the outcome, reasons for missingness and the missing fraction by treatment arm and cluster will be reported. Per protocol, the subjects are screened actively on their malaria status (the outcome) every four weeks.

In both cases, all the available data from the subject will be included in the primary and secondary analysis, without employing any specific missing data analysis techniques, due to the ignorability of the missing mechanisms. Missing baseline covariates (individual-level, household-level, and cluster-level) that are a part of the regression models for the outcome of interest will be imputed using simple hot-deck imputation methods if the missing fraction for the covariate is <5%. If the missing fraction for a covariable is ≥5%, appropriate multiple imputation approaches will be applied. If a non-ignorable portion of the subjects have missing values on a covariate (due to missing at random or missing completely at random), that covariate may be excluded in the model.

#### Additional analysis

We will perform the same analysis for three age subgroups (≤ 59 months old; 5 years old to 14 years old; 15 years old or older) to examine if the effects of Olyset®Plus ceiling net differ by age groups. In addition, we will perform other machine learning based approach such as causal forest and super learner to estimate the conditional average treatment effect.

#### Harms

Since the chance of having adverse event due to this intervention is very low based on the preceding study, all the details of unanticipated problems will be narratively reported, if any.

#### Statistical software

For all data handling and analysis, we will use R software version 4.3.2 (R Core Team (2021). R: A language and environment for statistical computing. R Foundation for Statistical Computing, Vienna, Austria. URL <https://www.R-project.org/>.)
